# Jinhua Qinggan granule, a Chinese herbal medicine against COVID-19, induces rapid changes in the plasma levels of IL-6 and IFN-γ

**DOI:** 10.1101/2020.06.08.20124453

**Authors:** Yasunari Kageyama, Koichi Aida, Kimihiko Kawauchi, Masafumi Morimoto, Tomoka Ebisui, Tetsu Akiyama, Tsutomu Nakamura

## Abstract

**Background:** Currently, effective vaccines or specific therapeutic agents against COVID-19 are not available. However, in China, traditional Chinese herbal medicines have provided therapeutic benefit to patients with COVID-19. Jinhua Qinggan granule (JHQGG) is a Chinese multi-herbal formula previously developed for the treatment of H1N1 influenza and has been encouraged for patients clinically suspected of COVID-19 during medical observation. However, the immunological mechanism for the efficacy of JHQGG has not been confirmed.

**Objectives:** We thus examined whether the administration of JHQGG affects hematological and immunological measures in healthy individuals.

**Method:** We enrolled 18 healthy volunteers, all of whom tested negative for antibodies to SARS-CoV-2. Peripheral blood samples were collected 1 h after oral administration of JHQGG and subjected to hematological, biochemical, and cytokine tests.

**Results:** JHQGG rapidly induced a significant decrease in the plasma level of IL-6 and an increase in the plasma level of IFN-γ.

**Conclusions:** Our finding suggests that the therapeutic efficacy of JHQGG against COVID-19 is, in part, associated with its rapid immunomodulatory activity.

## Introduction

There have been no specific vaccines or drugs proven to be clinically effective against COVID-19. However, in China, the majority of COVID-19 patients have been treated with a combination of traditional Chinese and modern Western medicine [1–3]. The use of several Chinese herbal formulas is encouraged for the treatment of COVID-19 in the latest version of the diagnosis and treatment protocol released by the National Health Commission of China. One of them is Jinhua Qinggan granule (JHQGG), which was formulated specifically for the symptoms of H1N1 influenza [4]. Previous preclinical studies showed that JHQGG reduces pulmonary lesions and mortality in mice infected with the H1N1 influenza virus [4]. A clinical study also demonstrated that JHQGG reduces the duration of fever and alleviates respiratory symptoms of patients with H1N1 influenza [4].

On the basis of its therapeutic efficacy for influenza, JHQGG has been recommended for patients clinically suspected of COVID-19 during medical observation. In a randomized controlled trial using mild cases in Wuhan, combined administration of JHQGG with Western medicine significantly ameliorated respiratory symptoms and relieved psychological anxiety compared with the administration of Western medicine alone [5–7]. However, the pharmacological mechanism underlying the efficacy of JHQGG has not been confirmed. We thus examined whether JHQGG administration affects hematological and immunological measures in healthy individuals.

## Materials and Methods

### Subjects

This is an open-label, single-arm study to obtain a clue to the pharmacological action of JHQGG. We enrolled a total of 18 healthy volunteers (5 males, 13 females; ages 22–58 years; mean age [SD], 33.8 [10.7] years), all of whom tested negative for IgM and IgG antibodies to SARS-CoV-2. Individuals were excluded if they had current infectious, inflammatory, or immune-related diseases.

### Administration of JHQGG

JHQGG was kindly provided by Hugh Wang, Juxiechang (Beijing) Pharmaceutical Co., Ltd. The subjects were instructed to take a packet (5 g) orally 40 min after lunch. This dose is known to be effective for the treatment of influenza [8]. Peripheral blood samples were obtained 1 h after the administration.

### Hematological, biochemical and cytokine analyses

Hematological and biochemical tests were outsourced to SRL, Inc. (Tokyo, Japan). Plasma cytokines were quantified using V-PLEX Proinflammatory Panel 1 Human Kit (Meso Scale Diagnostics) and Human IL-18 ELISA Kit (Abcam). All collected data (n = 18) were subjected to the statistical analysis using a two-tailed paired *t*-test.

## Results

Hematocrit and mean corpuscular volume changed marginally within the normal ranges (Table 1), but there were no significant differences in other measures of complete blood count and blood biochemistry between pre- and post-administration. Notably, in blood cytokine analysis, the plasma levels of IL-6 and IFN-γ were significantly decreased and increased, respectively, compared with those in pre-administration (IL-6, 2.75 vs. 1.63, 95% CI: −1.90 to −0.326, *P* = 0.00837; IFN-γ, 6.20 vs. 7.17, 95% CI: 0.193–1.73, *P* = 0.0172). The plasma IL-6 was decreased in 14 (77.8%) out of 18 subjects, whereas the plasma IFN-γ was increased in 13 (72.2%) out of 18 subjects (Fig. 1). We also found a relatively large decrease in the IL-18 level; however, the difference was not statistically significant (266 vs. 193, 95% CI: −154– 7.69, *P* = 0.0732).

**Table 1.** Hematological and cytokine changes in JHQGG-administered healthy individuals.

| Parameters | Pre-administration |  | Post-administration |  | Post – Pre | 95% CI | P-value |
| --- | --- | --- | --- | --- | --- | --- | --- |
|  | Mean | SD | Mean | SD |  |  |  |
| Complete blood count |  |  |  |  |  |  |  |
| Red blood cell count (x 10<sup>4</sup>/μl) | 448 | 50.3 | 445 | 52.1 | -2.94 | -7.58–1.69 | 0.198 |
| Hemoglobin (g/dl) | 13.4 | 1.79 | 13.3 | 1.81 | -0.100 | -0.229–0.0288 | 0.120 |
| Hematocrit (%) | 40.5 | 4.72 | 40.1 | 4.91 | -0.450 | **-0.851 to -0.0495** | **0.0298\*** |
| MCV (fl) | 90.7 | 6.50 | 90.3 | 6.28 | -0.456 | **-0.837 to -0.0744** | **0.0219\*** |
| MCH (pg) | 30.0 | 2.79 | 30.0 | 2.65 | -0.0444 | -0.257–0.168 | 0.665 |
| MCHC (%) | 33.0 | 1.31 | 33.2 | 1.24 | 0.139 | -0.0542–0.332 | 0.147 |
| White blood cell count (/μl) | 6170 | 797 | 6320 | 835 | 144 | -103–392 | 0.235 |
| Platelet count (x 10<sup>4</sup>/μl) | 25.5 | 4.84 | 25.6 | 5.17 | 0.0722 | -0.555–0.699 | 0.811 |
| Blood biochemistry |  |  |  |  |  |  |  |
| AST (U/l) | 17.1 | 3.95 | 17.1 | 3.72 | 0.00 | -0.482–0.482 | 1.00 |
| ALT (U/l) | 16.3 | 8.46 | 16.4 | 8.44 | 0.111 | -0.266–0.488 | 0.542 |
| γ-GT (U/l) | 26.9 | 44.6 | 27.2 | 44.8 | 0.278 | -0.0961–0.652 | 0.135 |
| LDH (U/l) | 150 | 30.0 | 147 | 26.0 | -3.00 | -7.67–1.67 | 0.193 |
| Albumin (g/dl) | 4.63 | 0.345 | 4.63 | 0.366 | 0.00 | -0.0782–0.0782 | 1.00 |
| Urea nitrogen (mg/dl) | 13.1 | 2.51 | 12.7 | 3.07 | -0.333 | -1.17–0.506 | 0.414 |
| HDL cholesterol (mg/dl) | 72.0 | 18.4 | 71.6 | 17.2 | -0.444 | -1.70–0.812 | 0.466 |
| LDL cholesterol (mg/dl) | 107 | 31.8 | 106 | 32.4 | -0.444 | -2.24–1.35 | 0.607 |
| Triglycerides (mg/dl) | 80.3 | 47.2 | 83.6 | 54.2 | 3.28 | -3.20–9.75 | 0.301 |
| CRP (mg/dl) | 0.102 | 0.200 | 0.106 | 0.209 | 0.00444 | -0.00127–0.0102 | 0.119 |
| Cytokines |  |  |  |  |  |  |  |
| IFN-γ (pg/ml) | 6.20 | 11.1 | 7.17 | 11.4 | 0.963 | **0.193–1.73** | **0.0172\*** |
| IL-12p70 (pg/ml) | 0.320 | 0.266 | 0.348 | 0.223 | 0.0279 | -0.0670–0.123 | 0.543 |
| IL-6 (pg/ml) | 2.75 | 3.02 | 1.63 | 1.82 | -1.11 | **-1.90 to -0.326** | **0.00837\*\*** |
| TNF-α (pg/ml) | 3.08 | 1.88 | 3.16 | 1.71 | 0.0849 | -0.152–0.322 | 0.460 |
| IL-1β (pg/ml) | 2.77 | 7.09 | 1.83 | 3.03 | -0.942 | -3.27–1.39 | 0.405 |
| IL-18 (pg/ml) | 266 | 273 | 193 | 160 | -73.3 | -154–7.69 | 0.0732 |
| IL-2 (pg/ml) | 0.532 | 0.763 | 0.643 | 0.795 | 0.111 | -0.0429–0.264 | 0.147 |
| IL-8 (pg/ml) | 734 | 808 | 1010 | 1420 | 274 | -123–671 | 0.163 |
| IL-10 (pg/ml) | 0.360 | 0.220 | 0.368 | 0.225 | 0.00792 | -0.0744–0.0902 | 0.841 |

MCV: mean corpuscular volume; MCH: mean corpuscular hemoglobin; MCHC: mean corpuscular hemoglobin concentration; AST: aspartate aminotransferase; ALT: alanine aminotransferase; γ-GT: γ-glutamyltransferase; LDH: lactate dehydrogenase; HDL: high-density lipoprotein; LDL: low-density lipoprotein; CRP: C-reactive protein; IFN: interferon; IL: interleukin; TNF-α: tumor necrosis factor-α; SD: standard deviation; CI: confidence interval. \* $P < 0.05$ , \*\* $P < 0.01$ .

**Fig 1.**
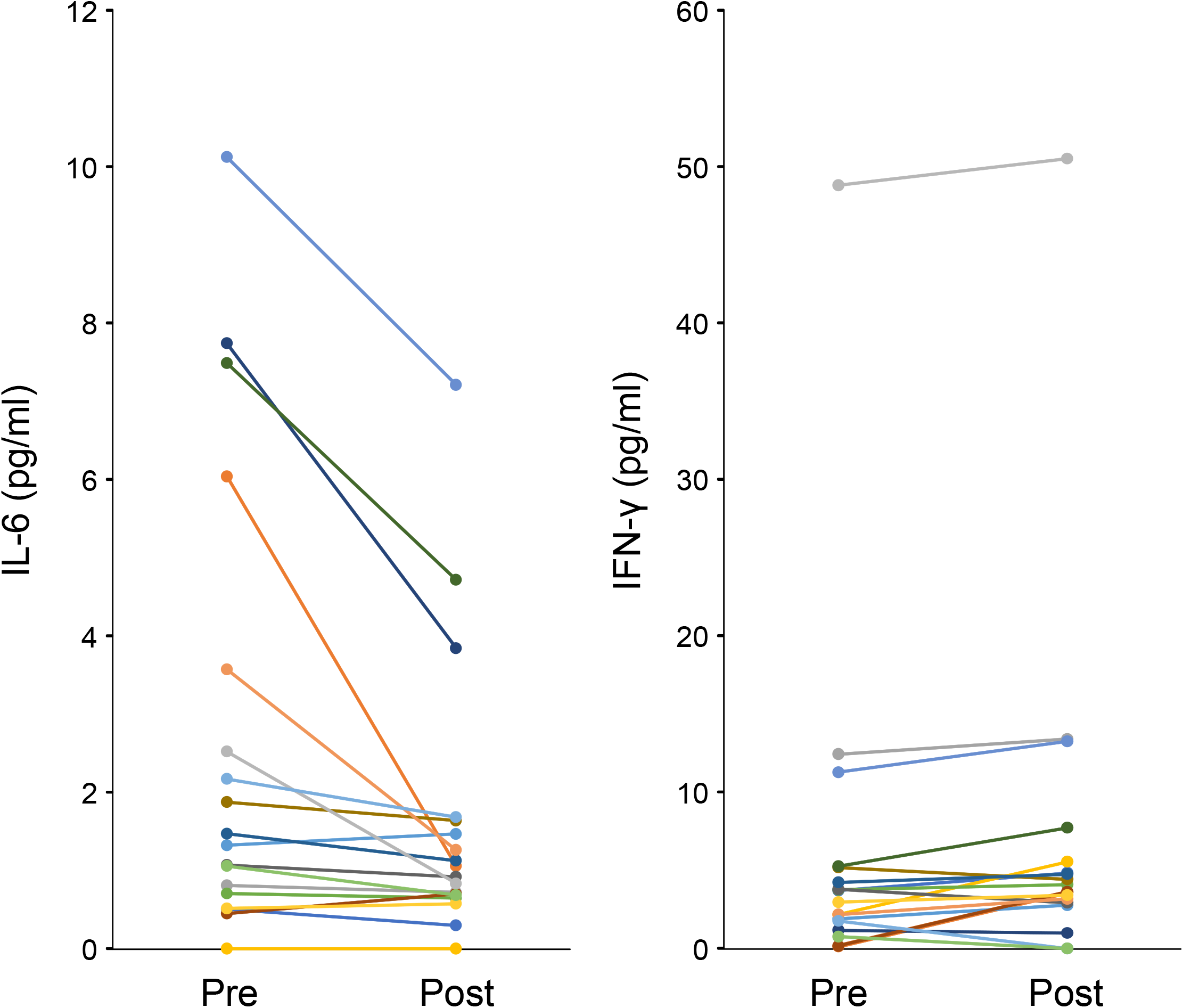
Changes in the plasma levels of IFN-γ and IL-6 before and after oral administration of JHQGG.

## Discussion

Dysregulated cytokine production is a hallmark of patients with COVID-19. Elevated blood levels of IL-2, soluble IL-2 receptor (sIL-2R), IL-4, IL-6, IL-10, and TNF-α have been observed especially in severe cases [9–12]. In particular, IL-6 plays pivotal roles in the exacerbation of COVID-19. Evidence is accumulating that an aberrant increase in IL-6 leads to complex immune dysregulation caused by SARS-CoV-2 infection. The blood IL-6 level is positively correlated with the severity and mortality in COVID-19 patients [13–15].

IFN-γ is a central mediator of antiviral immunity with an ability to directly interfere with viral replication. In addition, IFN-γ is indirectly involved in viral clearance through potentiating the action of IFN-α/β, activating Th1–dependent immune responses, and enhancing the MHC class I pathway [16]. Compared to healthy individuals, COVID-19 patients have significantly lower numbers of CD4^+^ T, CD8^+^ T, and NK cells with reduced capacity to produce IFN-γ, which causes the slightly decreased expression of IFN-γ [10,17]. Notably, the reduced cytotoxic potential of CD4^+^ T, CD8^+^ T, and NK cells is IL-6–dependent and severe COVID-19 cases have a higher IL-6/IFN-γ ratio than moderate cases [18].

In conclusion, JHQGG can down- and up-regulate the plasma levels of IL-6 and IFN-γ, respectively, immediately after oral administration. The rapid immunomodulatory effects of JHQGG may be able to remedy the immune dysregulation observed in COVID-19 patients and thus provide therapeutic benefit to mild to severe cases as well as asymptomatic or suspected cases.

## Data Availability

The data that support the findings of this study are available on request from the corresponding author, TN. The data are not publicly available due to their containing information that could compromise the privacy of research participants.

## Statements

## Acknowledgement

We thank Hugh Wang [Juxiechang (Beijing) Pharmaceutical Co., Ltd.] for generously providing us with JHQGG.

## Statement of Ethics

This study was carried out in accordance with The Code of Ethics of the World Medical Association (Declaration of Helsinki). All procedures were approved by the Ethics Committees of the Takanawa Clinic (approval number: 2020-2). A signed informed consent form was obtained from each participant prior to inclusion in this study. All experimental procedures and data analyses were conducted by investigators who were blinded to the subjects’ clinical information using a de-identified dataset. This study has been registered on University Hospital Medical Information Network-Clinical Trials Registry (UMIN-CTR) under the trial number UMIN000040407.

## Conflict of Interest Statement

Yasunari Kageyama, Koichi Aida, Kimihiko Kawauchi, Masafumi Morimoto, and Tomoka Ebisui are employees of Takanawa Clinic. Tetsu Akiyama and Tsutomu Nakamura have advisory roles in conducting clinical research in Takanawa Clinic and receive advisory fees from Takanawa Clinic.

## Funding Sources

This research did not receive any specific grant from funding agencies in the public, commercial, or not-for-profit sectors.

## Author Contributions

Yasunari Kageyama, Koichi Aida, Kimihiko Kawauchi, Masafumi Morimoto, and Tomoka Ebisui contributed to the conception and design of the study, contributed to data acquisition, analysis, and interpretation, and critically revised the manuscript for important intellectual content. Tetsu Akiyama contributed to the conception and design of the study and critically revised the manuscript for important intellectual content. Tsutomu Nakamura contributed to the conception and design of the study, contributed to data analysis and interpretation, and drafted the manuscript. All authors gave final approval of the submitted version of the manuscript and agree to be accountable for all aspects of the work.

## Notes

### Clinical Trial

UMIN000040407
We have prospectively registered our clinical trial on UMIN-CTR in May 15, 2020 prior to the start of the trial and has been opened the trial information to the public (https://upload.umin.ac.jp/cgi-open-bin/ctr/ctr_view.cgi?recptno=R000046108). We have described this information and the trial ID number (UMIN000040407) in our manuscript (refer to the Statement of Ethics section in page 4). UMIN-CTR (University Hospital Medical Information Network-Clinical Trials Registry) has been recognized by the ICMJE as an acceptable registry and is currently one of internationally recognized trial registries.

### Author Declarations

All procedures were approved by the Ethics Committees of the Takanawa Clinic (approval number: 2020-2).

